# Co-designing health-literate hand surgery care: qualitative priorities for patient education and patient-reported outcome feedback

**DOI:** 10.64898/2026.08.25.26361179

**Authors:** Ali Gholamrezaei, Dion Sandoz, Tanya Burgess, Brett McClelland

## Abstract

**Objective:** To identify patient, clinician, therapist and service priorities for a health-literacy intervention combining patient education with patient-reported outcome measure (PROM) feedback in routine hand surgery and hand therapy.

**Methods:** A qualitative co-design study was undertaken across public and private hand-care contexts in New South Wales, Australia. Twelve stakeholders participated: five consumers, three hand surgeons, one hand therapist and three administrative/managerial staff. Individual interviews plus a clinician group discussion were conducted. Data were collected in March 2026, audio-recorded, transcribed verbatim and de-identified. General inductive thematic analysis was undertaken in NVivo by one researcher, with final themes reviewed by co-investigators.

**Results:** Four themes guided intervention design: (1) providing information is not enough, it must be understood, retained and reinforced; (2) patients need a practical roadmap of diagnosis, treatment, recovery and rehabilitation; (3) education should be multimodal, reusable and adaptable to individual needs; and (4) PROMs should improve the clinical conversation rather than become another burden. Participants supported brief, accessible PROMs and visual feedback over time, but views differed on comparison with other patients because benchmarking could either reassure or create anxiety and unrealistic expectations.

**Conclusion:** Health-literate hand care requires more than readable leaflets. It requires repeated, practical and adaptable communication across the care pathway, with PROM feedback embedded in patient-clinician conversations.

**Practice implications:** Hand services should pair standardized core education with flexible delivery and use brief PROMs as conversation tools. Longitudinal displays may support monitoring and shared decisions, while group comparisons should be optional and carefully explained.

## 1. Introduction

Health literacy is shaped by both people’s capacities and the way health systems make information and services available, understandable and usable [1]. This is particularly relevant to surgery, where patients may need to understand a diagnosis, weigh treatment options, prepare for a procedure, follow restrictions, participate in rehabilitation and judge whether recovery is proceeding as expected. In hand surgery, limited health literacy is common and has been associated with differences in understanding, question-asking and clinical interaction [2–4].

Patient education is one obvious target, yet the problem is not solved by giving patients more written material. Hand-surgery resources have repeatedly been found to exceed recommended reading levels, and readability has shown limited improvement over time [5,6]. Health-disparities research in hand and upper-extremity care remains comparatively underdeveloped [7]. Visual strategies can improve comprehension and health-literacy-related outcomes, but effective communication also depends on timing, reinforcement, individual preference and the clinical environment in which information is used [8].

PROMs offer a complementary mechanism for making the patient perspective visible during care. They can capture symptoms, function and recovery in a standardized form and, when results are returned in an interpretable way, may help structure patient-clinician communication [11].

Hand-surgery research has highlighted both patient interest in meaningful outcome feedback and the lack of established processes for communicating individual PROM results at the point of care [12,13]. Orthopedic implementation studies likewise show that simply making PROM scores available does not ensure that clinicians will use them; workflow fit, interpretability and perceived clinical relevance matter [14,15]. Large-scale hand and wrist outcome systems demonstrate the feasibility of routine measurement and feedback loops, but translating these systems into locally usable communication tools remains a design challenge [16,17].

Co-design provides a way to develop these components with the people expected to use them rather than assuming that a single format or workflow will work for all. Co-design activities can legitimately include interviews and other forms of engagement when they contribute to iterative intervention development [20], and recent Patient Education and Counseling work has emphasized practical strategies for sustaining meaningful patient participation in intervention design [9,10]. We therefore undertook a qualitative co-design study to identify requirements for a multi-component Health Literacy Program for hand surgery and hand therapy. The study aimed to identify communication needs, educational priorities, requirements for meaningful PROM use and implementation considerations, and to translate these findings into concrete intervention components.

## 2. Methods

### 2.1. Design and setting

This qualitative co-design study formed Stage 1 of a three-year translational program to improve health literacy in hand surgery and hand therapy. The project involved public and private hand-care contexts in the Hunter New England region of New South Wales, Australia. The original protocol proposed three 3.5-hour mixed-stakeholder workshops. During recruitment, coordinating prolonged simultaneous attendance proved impractical and risked limiting consumer participation. The approved approach was therefore adapted to a flexible format using individual interviews and a clinician small-group discussion. The co-design purpose remained unchanged: to learn from lived and professional experience, identify intervention targets, explore early concepts and translate stakeholder input into intervention design.

### 2.2. Participants and recruitment

Purposive recruitment sought perspectives across lived experience, clinical roles, service roles and care settings. Twelve stakeholders participated: five consumers/patients, three hand surgeons, one hand therapist, and three administrative/managerial staff (a practice manager, administration assistant and office manager). Consumer participants were existing patients of participating hand surgery or therapy services who were invited through the clinical/research team. Clinicians and staff were invited because of their roles in hand-care delivery. Consumer experience included acute traumatic injuries, chronic/elective hand conditions including carpal tunnel syndrome and Dupuytren’s disease, repeated procedures and postoperative rehabilitation. Collectively, participants contributed experience spanning public and private care contexts. Detailed sociodemographic variables were not systematically collected in this co-design stage and are therefore not inferred or reported.

**Table 1.** Participant groups and contribution to co-design.

| Participant group | n | Data-collection format | Primary perspective |
| --- | --- | --- | --- |
| Consumers/patients | 5 | Individual interviews (~30 min) | Lived experience of information, surgery/therapy, recovery and PROM concepts |
| Hand surgeons | 3 | Clinician group discussion (~2 h) | Diagnosis/treatment communication, expectations, workflow and PROM use |
| Hand therapist | 1 | Clinician group discussion (~2 h) | Rehabilitation, self-management, communication and PROM use |
| Administrative/managerial staff | 3 | Individual interviews (~30 min) | Access, workflow and service implementation |

### 2.3. Data collection

Data collection occurred in March 2026. Interviews and the clinician discussion were conducted by a Research Associate who was not acting as the participants’ treating clinician. A semi-structured guide explored experiences of receiving and providing hand-care information; understanding of diagnosis, treatment and recovery; rehabilitation and self-management; preferred information formats; experiences of monitoring progress; and views on collecting and using PROMs in routine care. The guide was not formally pilot tested.

Individual interviews were conducted using a mix of in-person and remote formats and typically lasted approximately 30 minutes. The group discussion with three hand surgeons and one hand therapist lasted approximately two hours. All sessions were audio-recorded with consent, transcribed verbatim, checked and de-identified before analysis. Early concepts for PROM collection and visualization were shown or described to participants where relevant, allowing discussion of score interpretation, color cues, longitudinal graphs and comparison with other patients.

### 2.4. Analysis

A general inductive approach was used to develop themes from the data rather than coding against a predefined theoretical framework [18]. Transcripts were managed and coded in NVivo. The interviewer conducted the initial coding, compared recurrent ideas across consumer, clinical and service perspectives, grouped related codes into candidate themes and iteratively refined the thematic structure around implications for intervention design. The final themes were reviewed by co-investigators for coherence with the source data and the purpose of the co-design study. No formal claim of data saturation was made.

### 2.5. Ethics and reporting

Ethics approval was obtained from the Hunter New England Local Health District Human Research Ethics Committee (2025/ETH01957; approved 16 September 2025, amendment approved 30 September 2025), with site-specific authorizations for participating services. Participants provided informed consent. Reporting was guided by the Consolidated Criteria for Reporting Qualitative Research (COREQ) [19]; the completed checklist is provided as supplementary material.

## 3. Results

The adapted format enabled participation from all 12 stakeholders and generated a set of convergent but not uniformly identical priorities. Four overarching themes captured the findings. Importantly, participants did not simply ask for “more information”. They described a communication system in which information is reinforced at appropriate times, tailored to practical needs, available in multiple formats and connected to clinically meaningful feedback about recovery.

### 3.1. Providing information is not enough: it must be understood, retained and reinforced

Consumers often described good care when clinicians used ordinary language, gave consistent explanations and were available to clarify questions. One consumer contrasted technical terminology with the plain-language explanation they received:

“It was all clear and easy to understand … they explained it in common terms … everyone was on the same page.” — Consumer 5

The clinician group, however, emphasized that information provision and information retention were different problems. Patients could receive detailed verbal explanations, written consent information and postoperative instructions, yet later appear to encounter the same information as if for the first time. One clinician summarized this directly:

“I think retention of information is a problem.” — Clinician group

Participants linked poor retention to timing and cognitive load. Surgical decisions can occur when patients are anxious, in pain or processing several practical consequences at once. This suggested that the intervention should not rely on a single information event. Core messages should be accessible for later review and reinforced across consultation, surgery and rehabilitation. Consistency across professionals was also important because contradictory explanations could add avoidable uncertainty, particularly in trauma pathways involving multiple services.

### 3.2. Patients need a practical roadmap of what happens next

Participants wanted information to connect diagnosis with what it meant for everyday life. This included basic anatomy and pathology, treatment choices, why a particular option was recommended, likely restrictions, therapy requirements, expected milestones and uncertainty in the pace of recovery. Consumers frequently described the postoperative period as a time when practical questions became more salient than they had been before surgery.

“It’s when your head starts to clear … and that’s when you need the most support.” — Consumer 1

This consumer described the effects of hand injury on work, family responsibilities, cooking and mental wellbeing. Another emphasized the importance of realistic expectations:

“It’s not necessarily a couple of weeks problem … it can be a long-term problem with lots of therapy.” — Consumer 2

Clinicians similarly described difficulty communicating the natural variation around recovery. Generic statements such as an average return-to-work time could be interpreted by patients as a promise rather than a range. The design implication was therefore a “recovery roadmap” that explains what is expected, what varies between individuals, what patients can do to support recovery and what symptoms or changes should trigger contact with the care team. Postoperative education was viewed as part of the treatment itself rather than an administrative handout.

### 3.3. Health-literate education should be multimodal, reusable and adaptable

Preferences varied markedly. Some consumers valued verbal explanation most; others wanted diagrams, written instructions, links to trusted websites or more detailed scientific information. A participant with Dupuytren’s disease described the value of layered information:

“Having the written word as well as diagrams and maybe even a glossary … might be really useful.” — Consumer 3

Visual explanations were particularly valued for anatomy, the location of pathology and rehabilitation exercises. Reusable resources were also important because instructions may need to be revisited after the consultation. Participants suggested short videos for exercises or typical recovery trajectories, while recognizing that video would not suit everyone. Digital access through a phone or computer was convenient for many participants, but paper, email or in-clinic options remained necessary for people who preferred them or lacked digital access.

Clinicians strongly supported a standardized core but were wary of one-size-fits-all education. Existing materials were characterized as “very generic”. Clinicians described changing their language and level of detail according to the patient’s apparent understanding and goals, while acknowledging that this judgement is imperfect. The intervention therefore needed a stable set of core messages with enough flexibility to add detail, visuals or tailored advice without requiring clinicians to recreate education from scratch for every patient.

### 3.4. PROMs should improve the clinical conversation, not become another burden

Participants generally supported PROMs when the purpose was clear and completion burden was low. One consumer suggested that a brief pre-visit questionnaire could capture a symptom that might otherwise be forgotten by the time of the appointment, and considered approximately five to ten minutes acceptable. Another participant cautioned that questionnaires should not be treated as a substitute for clinical dialogue:

“Questionnaires are not fluid … the conversation you have with the therapist is dynamic.” — Consumer 4

This distinction shaped the proposed role of PROMs: they should prompt and focus conversation rather than determine treatment. Participants preferred outputs that made changes over time visible. A consumer who had actively tracked finger measurements suggested that a simple chart could make recorded progress easier to share, and another responded positively to color-coded symptom categories and longitudinal displays.

Views on benchmarking against other patients were deliberately divergent. One consumer valued seeing how they were tracking and said, “I really like the selected patient versus others”, but also thought such information should be optional because poorer-than-average recovery could increase stress. Another rejected comparison more strongly: “Because everyone heals different.” This led to a design requirement that individual longitudinal change should be central, while group-reference information should be used selectively, with explanation of uncertainty and individual variation rather than presented as a normative recovery target.

**Table 2.**
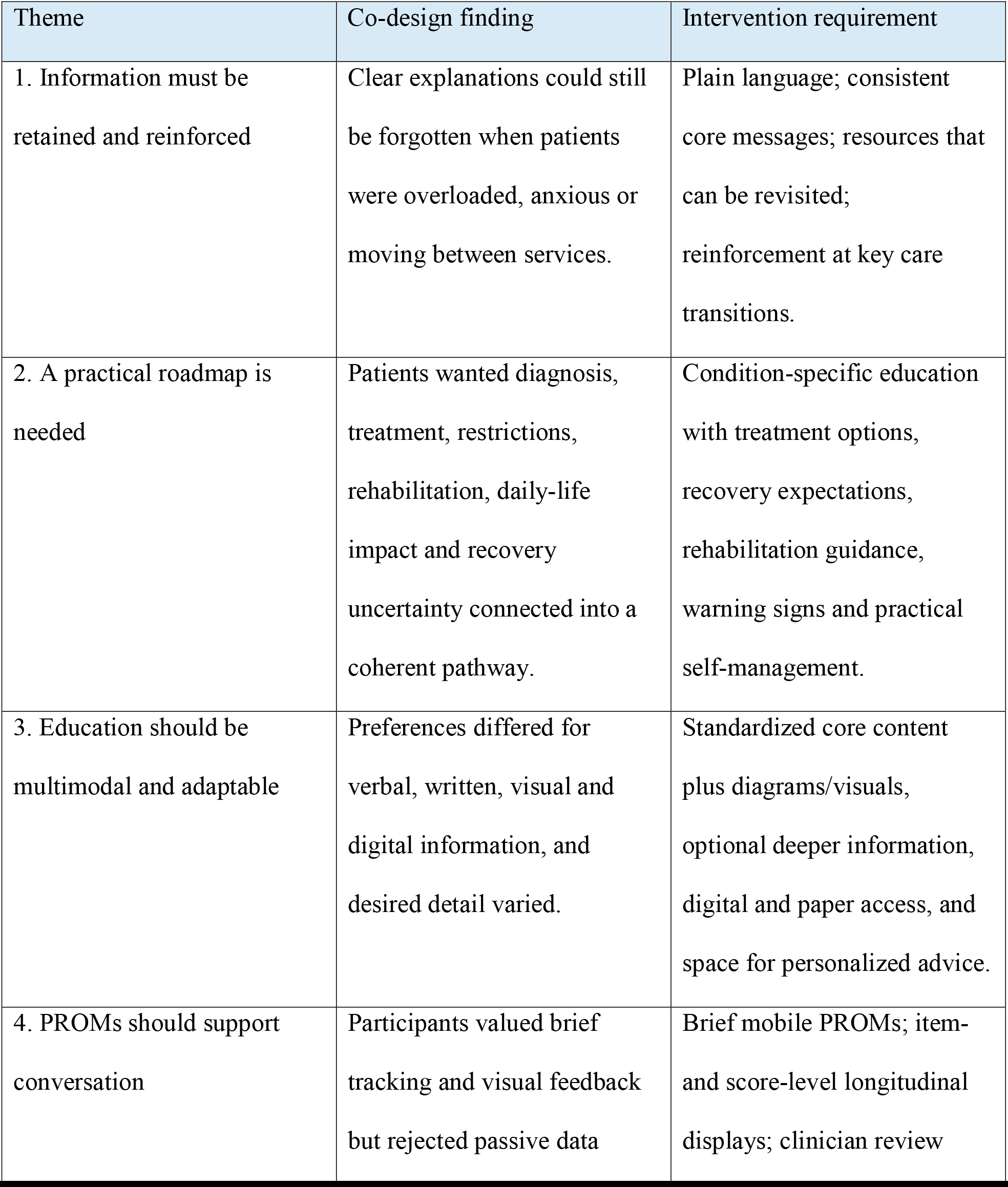
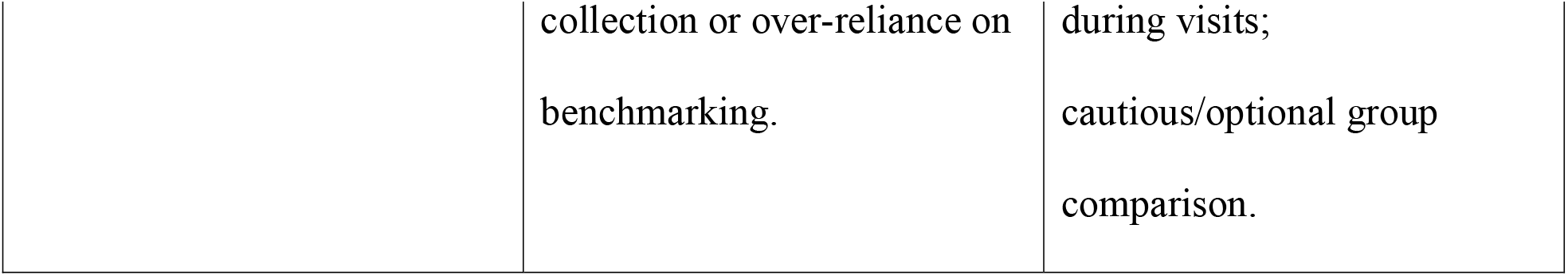
Themes and resulting intervention requirements.

### 3.5. Translation of findings into intervention design

The themes were translated into two linked intervention components. First, standardized condition-specific patient education was developed around plain language, simple visuals, treatment expectations, self-management, postoperative guidance, warning signs, questions for shared decision-making and a short explanation of how PROMs would be used. Second, a clinician-facing PROM dashboard was developed to display longitudinal HAND-Q symptom and function information at both summary-score and item level. The dashboard uses simple severity cues and time-series displays so that PROM data can be reviewed during a consultation rather than stored as passive research data. Figure 1 illustrates the resulting design using dummy/test data; it is presented as an output of the co-design translation process, not as evidence of effectiveness.

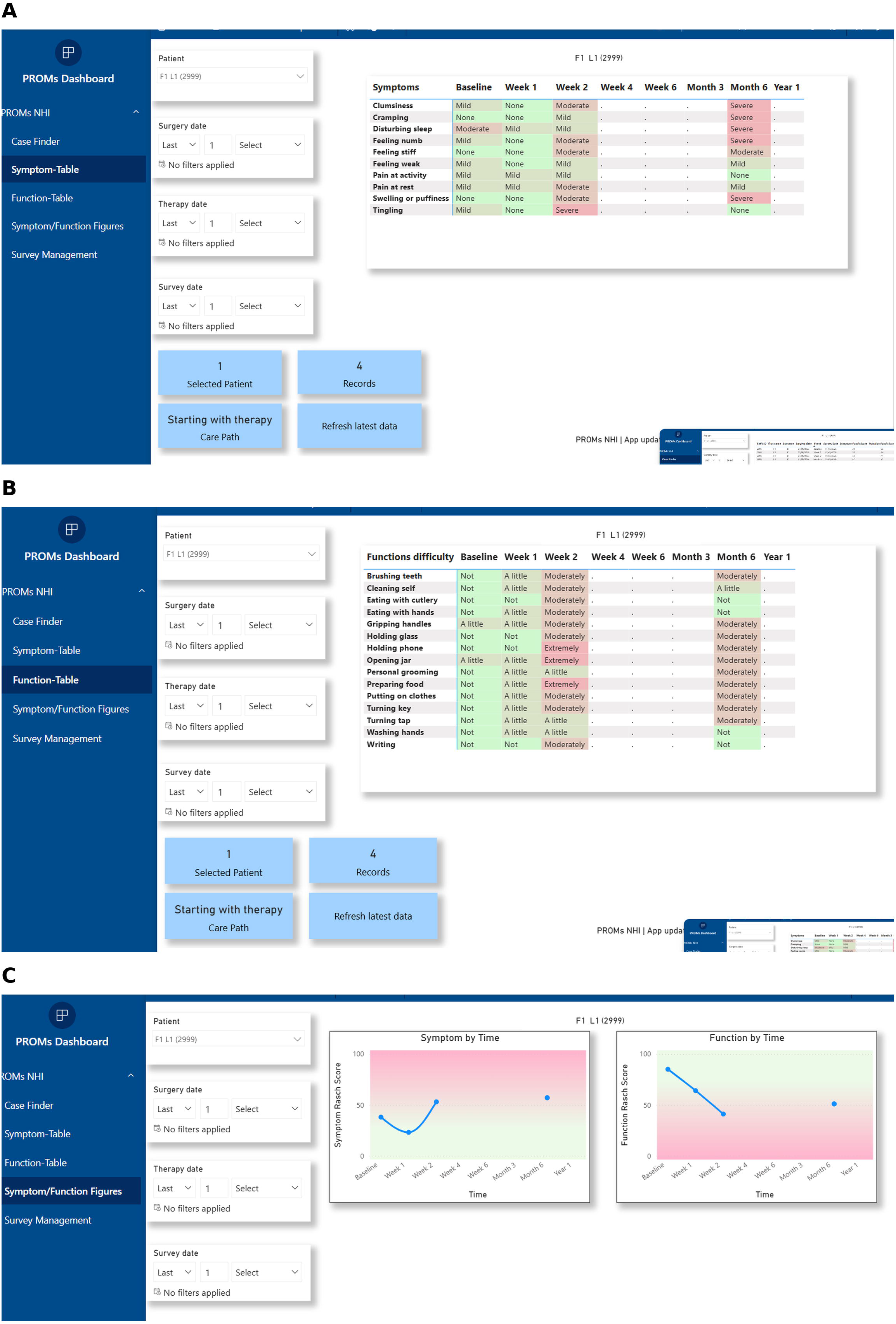

## 4. Discussion

This study identified four practical requirements for health-literate hand care: information must be reinforced rather than merely delivered; patients need a realistic roadmap through treatment and recovery; education should be multimodal and adaptable; and PROMs should serve the clinical conversation rather than compete with it. The value of the findings lies in the connection between these requirements. A readable leaflet alone does not solve the problem if it is given at the wrong time, cannot be revisited, does not explain recovery, or is disconnected from subsequent clinical conversations. Conversely, a sophisticated PROM platform adds little if scores are difficult to interpret or are not actively used during care.

The findings support an organizational view of health literacy [1]. Consumers in this study often had the capacity to seek information, ask questions or use digital resources, yet still described circumstances in which timing, information load, access or uncertainty affected what could be used. Clinicians described similar problems from the service side, particularly inconsistency and poor retention across the care pathway. This shifts the intervention target from “fixing” an individual patient’s literacy to designing a system that reduces unnecessary communication demands. It also aligns with evidence that hand-surgery information is frequently written at a level that may exclude patients with lower literacy [5,6] and with broader evidence supporting visual approaches to make health information more usable [8].

A second contribution is the emphasis on recovery as an information need in its own right. Consumers were not only interested in the mechanics of surgery; they wanted to know what the next days, weeks and months might mean for hand use, work, caring responsibilities, therapy and uncertainty. Clinicians described the same problem from the opposite direction: patients may interpret an average recovery time as a fixed promise. The intervention response is therefore not simply to add a timeline but to communicate a range, explain variability and pair expected milestones with practical actions and escalation advice. This resembles recent co-design work in Patient Education and Counseling in which communication tools were shaped around transitions and continuity rather than information content alone [10].

The strong preference for multimodal and layered information also argues against choosing a single “best” medium. Participants differed in whether they preferred verbal explanation, diagrams, written information, video or more detailed sources. Clinicians likewise described tailoring language and detail during consultations. Co-design guidance emphasizes that meaningful participation requires attention to context, roles and methods rather than adherence to one engagement format [9,20]. Our pragmatic shift from fixed workshops to individual interviews and a clinician discussion is consistent with this principle: interviews are recognized co-design activities when participant input is iteratively translated into the design [20]. The adaptation also reduced a practical participation barrier created by asking consumers and clinicians to attend several long sessions simultaneously.

The PROM findings are particularly relevant to current hand-surgery practice. Patients in previous hand-surgery research have reported that PROMs can be valuable when they capture personally important goals and changes [12]. At the same time, the Hand Surgery Quality Consortium concluded that stronger process guidance is still needed for routine collection and communication of individual scores [13]. Our participants supplied concrete design requirements: keep questionnaires brief, show change over time, make scores interpretable, and review results in the clinical encounter. The concern that questionnaires are “static” compared with a dynamic conversation reinforces the realist-synthesis finding that PROMs influence care through the way information is interpreted and acted on, rather than through measurement alone [11]. Clinician-focused orthopedic research similarly shows that availability without workflow integration produces low use [14].

The divergent views on benchmarking deserve specific attention. Routine hand outcome systems have successfully used comparison with group trajectories to contextualize recovery and support shared decision-making [17]. In our study, one consumer actively wanted this comparison while others worried that it could create anxiety or an inaccurate expectation of when they “should” recover. This tension is consistent with calls for equitable and careful implementation of PROM feedback, particularly when individual outcomes are influenced by clinical, psychological and social factors [15]. For the current intervention, the design consequence is to prioritize a patient’s own trajectory and make group comparison optional, contextual and clinician-mediated rather than an automatically displayed target.

The study has several strengths. It combined consumer, surgical, therapy and service perspectives; included experience across acute and chronic conditions and public/private care contexts; sought negative and divergent views rather than forcing consensus; and translated qualitative findings into tangible educational and digital design requirements. The clinician-facing dashboard provides a visible example of how stakeholder comments about brevity, color cues, longitudinal feedback and conversation support were operationalized. This translational link is important because co-design reports are more useful when they describe what changed as a result of participation [9].

Limitations should also be recognized. The sample was small and drawn from one Australian region, so the findings are intended to inform intervention design rather than estimate the frequency of views. Detailed sociodemographic characteristics were not systematically collected, limiting assessment of transferability across literacy, language and socioeconomic groups. Initial coding was undertaken by one researcher, although themes were subsequently reviewed by co-investigators. Different stakeholder groups contributed through different formats, so consumers and clinicians did not always generate ideas together in the same session. No formal saturation assessment was performed. Finally, the dashboard and educational resources described here are intervention outputs; their feasibility, acceptability, accessibility and effectiveness require prospective evaluation and should not be inferred from this co-design study.

## 5. Conclusion

Health-literate hand care requires more than simplifying written information. Stakeholders identified a need for clear messages that are reinforced over time, a practical roadmap through recovery, multimodal resources that can be adapted to individual needs, and PROM feedback that strengthens rather than replaces patient-clinician communication. Translating these requirements into standardized patient education and a clinician-facing PROM dashboard provides a testable model for the next phase of implementation and evaluation.

## Practice implications

Hand-care services should provide a standardized core explanation of diagnosis, treatment, recovery and rehabilitation, but allow patients to access information through verbal, written, visual and digital formats according to preference and need. PROMs should be brief, returned in an interpretable longitudinal form and actively discussed during care. Group-reference trajectories may be useful for some patients, but should be optional and accompanied by explanation of individual variability.

## Declarations

### Ethics approval

Hunter New England Local Health District Human Research Ethics Committee, 2025/ETH01957.

### Data availability

Individual-level interview transcripts are not publicly available because participant consent and ethics approval do not provide for public sharing. De-identified aggregated findings are reported in this article.

## Acknowledgements

We thank the consumers, clinicians, therapist and service staff who contributed to the co-design process.

